# Vascular Phenotyping in Parkinson’s Disease: Diabetes Mellitus Operationalizes a Microvascular Metabolic Syndrome Cluster Across PPMI Diagnostic Cohorts

**DOI:** 10.64898/2026.06.09.26355285

**Authors:** Alexander Belnavis, Shannon Y. Chiu, Kewei Chen, Roland J. Thorpe, Edward Ofori, the Parkinson Precision Medicine Initiative

## Abstract

**Background:** Diabetes mellitus elevates Parkinson’s disease (PD) risk, via hypothesized cerebrovascular mediation. Whether the diabetes/prediabetes vascular-risk phenotype concentrates in cardiometabolic risk or macrovascular events across prodromal and clinically diagnosed PD remains unresolved.

**Objectives:** To quantify the vascular-risk burden associated with diabetes/prediabetes across the PPMI diagnostic cohorts to test whether this association differs by cohort.

**Methods:** Cross-sectional analysis of 413 PPMI participants (76 healthy controls, 145 prodromal PD, 192 clinically diagnosed PD) examined diabetes/prediabetes (n = 73) and seven vascular risk factors. The Vascular Burden Score (0 to 7) was a priori partitioned into microvascular and macrovascular sub-scores. Modified Poisson regression estimated adjusted prevalence ratios (aPR), adjusted for age, sex, and body mass index. A cohort-by-diabetes interaction tested cross-cohort consistency. Sensitivity analyses incorporated nigral diffusion tensor imaging (PD-risk biomarker) and FreeSurfer white matter hypointensity volume (cerebrovascular marker).

**Results:** Diabetes/prediabetes elevated Vascular Burden Score (β = 0.53, 95% CI 0.29 to 0.77, p < 0.001) versus non-diabetic participants, with a non-significant cohort-by-diabetes interaction (F = 0.29, p = 0.747). Three microvascular factors survived false discovery rate correction: obesity (aPR 2.28), hypertension (aPR 1.60), and hyperlipidemia (aPR 1.45). Macrovascular events showed no diabetic amplification (β = −0.06, p = 0.25). In the imaging-phenotyped subset, Vascular Burden Score components contributed classifier variance distinct from nigral microstructure.

**Conclusions:** Diabetes/prediabetes operationalize a microvascular cluster stable across prodromal and idiopathic PD. Cardiometabolic phenotyping may complement established PD-risk biomarkers (dopamine transporter SPECT, nigral diffusion), pending longitudinal validation linking vascular phenotype to dopaminergic markers.

## Introduction

A central challenge in Parkinson’s disease (PD) is detection of risk before clinical symptom onset. PD is defined by degeneration of dopaminergic neurons in the substantia nigra, but motor manifestations are preceded by years of non-motor symptoms including autonomic dysfunction, sleep disturbances, and cognitive change.^1-4^ Among the modifiable contributors to this prodromal phase, vascular risk factors including diabetes mellitus, hypertension, and dyslipidemia drive cerebrovascular small vessel disease that may degrade the neurovascular substrate supporting nigrostriatal integrity well before motor onset.^5,6^

Cerebral small vessel disease, characterized by white matter hyperintensities, lacunar infarcts, microbleeds, and enlarged perivascular spaces, compromises blood-brain barrier integrity, disrupts cerebral autoregulation, and degrades the structural environment supporting basal ganglia circuits.^5,7^ Mendelian randomization analyses have established that diabetes mellitus, elevated blood pressure, and adiposity exert causal effects on small vessel disease burden, identifying these as upstream targets rather than downstream consequences of shared aging biology.^8^ Convergent neurobiology evidence implicates blood-brain barrier compromise and neurovascular unit dysfunction as shared mechanisms across neurodegenerative disorders.^12^ Longitudinal studies show small vessel disease progression independently increases risk of incident parkinsonism, positioning cerebrovascular substrate degradation as a precursor to clinical PD onset.^9-10^ When small vessel disease coexists with PD, it accelerates motor and cognitive decline with severity correlating with Hoehn and Yahr staging.^6,11^

Beyond conventional structural imaging, diffusion tensor (DTI) quantifies microstructural integrity within deep brain regions including the substantia nigra. Free-water and diffusion tensor metrics in the substantia nigra are sensitive to early nigrostriatal degeneration and have been advanced as candidate imaging biomarkers of dopaminergic neurodegeneration in PD.^28^ T1-derived white matter hypointensity volume provides a complementary structural surrogate for cerebrovascular burden from routine clinical imaging.^39^

Approximately 35.4 million individuals in the United States are living with diabetes mellitus.^13^ Diabetes mellitus is a metabolic disease defined by inappropriately elevated blood glucose levels, with type 2 diabetes mellitus characterized by insulin resistance and a relative functional insulin deficit.^14^ Type 2 diabetes mellitus is characterized by insulin resistance which in turn has linked the metabolic disease to neurodegenerative conditions, with prior research showing 5.6% of PD patients have newly diagnosed type 2 diabetes mellitus and 26% have insulin resistance.^15^ Epidemiological meta-analysis demonstrates individuals with diabetes mellitus have a 27% increased risk of developing PD and individuals with prediabetes a 4% increased risk.^16^

Despite the convergent evidence linking diabetes mellitus to cerebrovascular pathology and elevated PD risk, the diabetes mellitus and prediabetes associated vascular phenotype has not been quantified within a single PD biomarker cohort that includes healthy controls, individuals with prodromal PD, and clinically diagnosed PD participants. This gap leaves unresolved whether the vascular phenotype is detectable before clinical PD onset, and whether it operates through cumulative microvascular pathology or through discrete atherothrombotic and prothrombotic events. The present cross-sectional analysis of Parkinson’s Precision Medicine Initiative (PPMI) participants addresses this gap through three aims: characterize the diabetes mellitus and prediabetes associated vascular phenotype using a composite Vascular Burden Score (VBS);^36^ test whether the phenotype operates through microvascular versus macrovascular pathways; and evaluate whether the phenotype is stable across PPMI diagnostic cohorts. Sensitivity analyses test association with structural cerebrovascular damage on FreeSurfer-derived T1 white matter hypointensity volume (Figure 3D); an exploratory random forest classification analysis evaluating Vascular Burden Score components against nigral diffusion microstructure is presented (see Supplementary Analysis S2).

## Methods

This cross-sectional analysis used baseline data from PPMI, an international multicenter observational study of PD biomarkers.^17^ Participants in the analytic sample were enrolled in PPMI 1.0 between June 2010 and June 2019; prodromal and clinically diagnosed PD enrollment under PPMI 1.0 required dopamine transporter SPECT (DaT scan) confirmation of dopaminergic deficit. All participants provided written informed consent. Participants were eligible for inclusion if they had complete baseline data for diabetes mellitus/prediabetes status, all seven vascular risk factor indicators, and the demographic covariates (age, sex, body mass index). Prodromal status was defined using the Movement Disorder Society research criteria.^18^ Participant selection is shown in Figure 1. Detailed information about PPMI study design, inclusion/exclusion criteria, informed consent, and demographic data, can be found on the website (https://www.ppmi-info.org/). Per PPMI publication policy, this manuscript was submitted to the PPMI Data Use and Publications Committee for administrative compliance review prior to journal submission.

**Figure 1.**
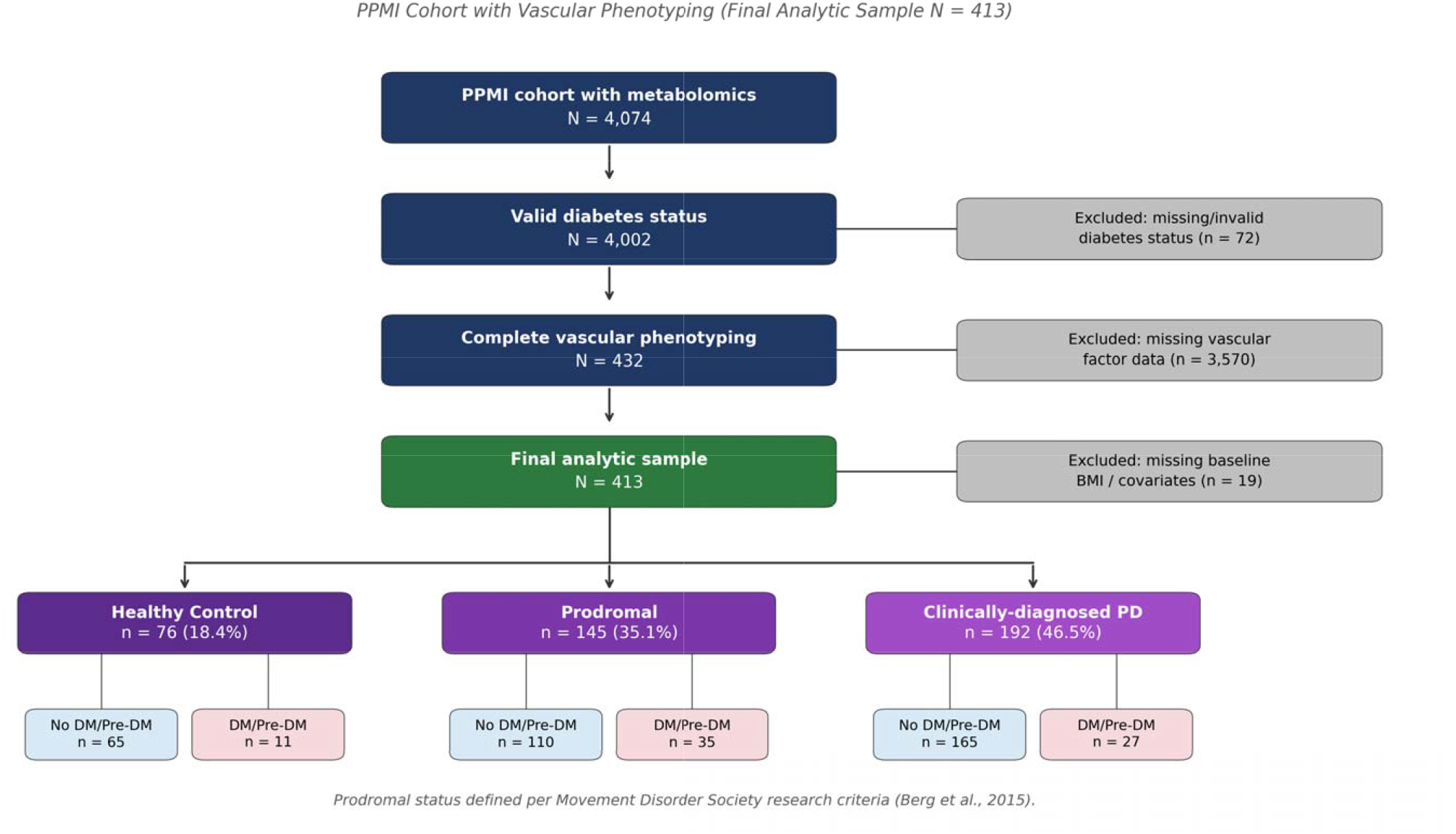
Participant Selection Flowchart. Participant selection from the PPMI metabolomics cohort (N = 4,074) to the final analytic sample (N = 413). Exclusion criteria applied sequentially: missing diabetes mellitus/prediabetes status, missing vascular factor data, missing body mass index, and incomplete demographic covariates. Final cohort composition: 76 healthy controls, 145 prodromal individuals (per Movement Disorder Society research criteria), and 192 clinically diagnosed Parkinson’s disease participants. The diabetes mellitus/prediabetes group comprised 73 participants (17.7%).

Diabetes mellitus/prediabetes status was ascertained through systematic regular expression search of the PPMI Medical Conditions Log for diagnostic terms including diabetes mellitus, type 2 diabetes mellitus, T2DM, prediabetes, and impaired glucose tolerance, with manual review of ambiguous entries. Participants with any qualifying entry were coded as positive (DM_Group = 1); all others were coded as negative (DM_Group = 0).

The composite Vascular Burden Score (VBS) was constructed as the count of seven binary vascular risk factor indicators (range 0 to 7), building on composite cardiovascular risk scoring approaches established in cognitive aging research^36^: hypertension, hyperlipidemia, obstructive sleep apnea, obesity (body mass index ≥ 30 kg/m^2^ per WHO criteria^19^), stroke or transient ischemic attack, coronary artery disease, and atrial fibrillation. The first six factors were ascertained from the PPMI Medical Conditions Log; obesity was operationalized as a binary indicator (yes/no) by dichotomizing baseline body mass index at the WHO cutoff of 30 kg/m^2^. The Vascular Burden Score was partitioned a priori into a four-component microvascular sub-score (hypertension, hyperlipidemia, sleep apnea, obesity; range 0 to 4) capturing factors with established mechanistic links to small vessel pathology and metabolic syndrome,^20,23^ and a three-component macrovascular sub-score (stroke or transient ischemic attack, coronary artery disease, atrial fibrillation; range 0 to 3) capturing discrete atherothrombotic and prothrombotic events not included in standard metabolic syndrome definitions. We use the term ‘microvascular’ to denote vascular-risk factors with established links to small vessel disease and metabolic syndrome biology, rather than direct imaging-confirmed cerebral microvascular injury.

Primary models adjusted for age (continuous, years), sex (binary), and body mass index (continuous, kg/m^2^). For models in which obesity was the dependent vascular factor, body mass index was omitted because obesity was derived directly from body mass index. Body mass index was also omitted from the microvascular sub-score model because obesity is one of the score components.

### Statistical Analysis

Sample characteristics were compared between diabetes mellitus/prediabetes positive and negative groups using independent samples t-tests for continuous variables and chi-square tests for categorical variables. Per-factor associations between diabetes mellitus/prediabetes status and each of the seven vascular factors were tested using modified Poisson regression with robust variance estimation.^21,30-31^ Models were adjusted for age, sex, and body mass index. Benjamini-Hochberg false discovery rate correction was applied across the seven factors to control multiple comparisons.^22^

The independent effect of diabetes mellitus/prediabetes on three composite outcomes was tested through multivariable linear regression: total Vascular Burden Score (Model A), microvascular sub-score (Model B, body mass index omitted), and macrovascular sub-score (Model C). Because Vascular Burden Score is a bounded count outcome, count-model sensitivity analyses using Poisson or negative binomial regression were performed to confirm the robustness of the linear-model inference. Whether the diabetes mellitus/prediabetes effect on Vascular Burden Score varied across PPMI clinical cohort was tested through a two-way univariate analysis of variance with cohort (healthy control, prodromal, PD) and diabetes mellitus/prediabetes as factors, adjusted for age, sex, and body mass index. Within-cohort effect sizes were quantified as Cohen’s d with Hedges’ small-sample correction.

PPMI diffusion tensor imaging (DTI) data were preprocessed by the PPMI Imaging Core using the Center for Imaging of Neurodegenerative Diseases pipeline,^29^ which performs eddy-current and motion correction, mutual-information registration of DWI to T1- and T2-weighted structural MRI, and nonlinear EPI distortion correction. Standard DTI metrics were derived in the corrected EPI space for six substantia nigra sub-regions and averaged within each participant: fractional anisotropy (FA, Equation 1),

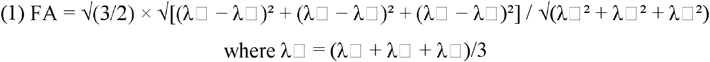

and mean diffusivity (MD, Equation 2) per the CIND pipeline definitions,^29^

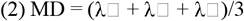

and radial diffusivity (RD, Equation 3) computed from the pipeline-derived eigenvalues following standard tensor decomposition conventions.^34^

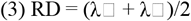

These three metrics align with the diffusion MRI features previously associated with PD-related nigral microstructural change.^28^

Two sensitivity analyses were performed to assess the robustness of the primary findings against alternative confounders and to characterize Vascular Burden Score relative to a complementary structural imaging biomarker. The first restricted analysis to clinically diagnosed PD participants (n = 192) with adjustment for years since diagnosis, testing whether the observed phenotype was driven by PD chronicity. The second regressed log-transformed normalized FreeSurfer 7-derived T1 white matter hypointensity volume on Vascular Burden Score, adjusted for age, sex, and body mass index, in the subset of 318 participants with available FreeSurfer 7 ASEG output.^35,39^ An additional exploratory random forest analysis evaluating Vascular Burden Score components against nigral diffusion metrics (FA, MD, RD) in the imaging subset (n = 50) is reported in Supplementary Analysis S2 with detailed results in Supplementary Table S6.

Primary and sensitivity analyses were conducted in IBM SPSS Statistics version 30. Modified Poisson regression and the supplementary random forest analyses were performed in Python 3.11 (scikit-learn 1.5). Statistical significance was set at α = 0.05 (two-sided). Analysis code is available at the repository indicated in the Data Availability Statement.

## Results

The final analytic sample comprised 413 PPMI participants (mean age 71.7 ± 7.9 years, 47.0% female), distributed across three cohorts: 76 healthy controls (18.4%), 145 individuals with prodromal PD (35.1%), and 192 clinically diagnosed PD participants (46.5%). Diabetes mellitus/prediabetes was present in 73 participants (17.7%) and absent in 340 (82.3%). Mean composite Vascular Burden Score was 1.97 ± 1.13 in the diabetes mellitus/prediabetes group and 1.17 ± 1.10 in the non-diabetic group (Table 1). Detailed baseline clinical characterization stratified by cohort and diabetes mellitus/prediabetes status, including MDS-UPDRS scores, Hoehn & Yahr stage, MoCA, and LRRK2 and GBA variant frequencies was compiled (see Supplementary Table S1).^40,41^

**Table 1.**
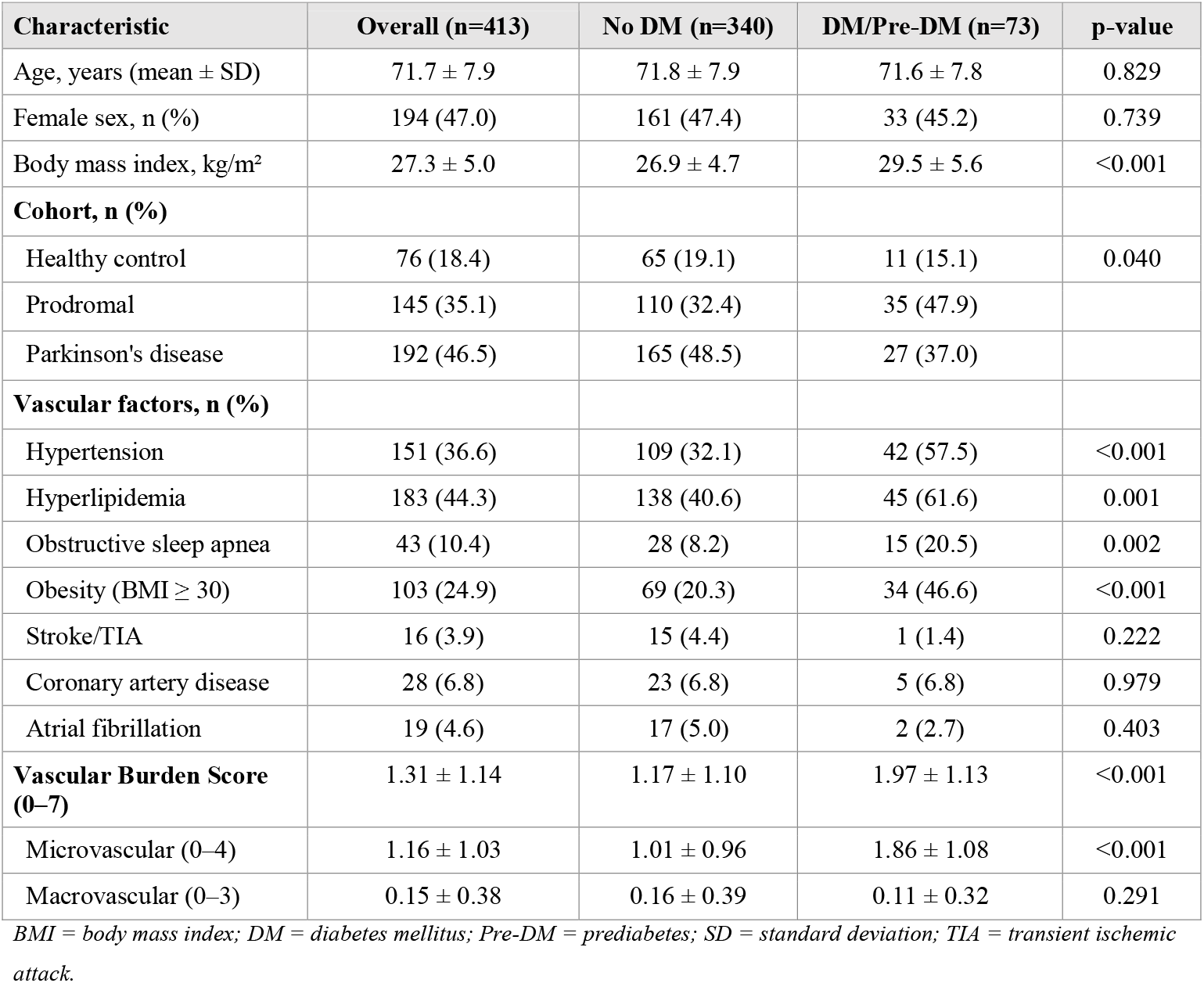
Distribution of sample characteristics and vascular factor prevalence by diabetes mellitus/prediabetes status of 413 participants in PPMI database. Demographics, body mass index, and prevalence of each vascular risk factor are shown by diabetes mellitus/prediabetes group. Continuous variables (age, body mass index) compared via independent samples t-tests; categorical variables compared via chi-square tests. Composite Vascular Burden Score (range 0 to 7) and microvascular (range 0 to 4) and macrovascular (range 0 to 3) sub-scores presented as mean ± standard deviation.

| Characteristic | Overall (n=413) | No DM (n=340) | DM/Pre-DM (n=73) | p-value |
| --- | --- | --- | --- | --- |
| Age, years (mean ± SD) | 71.7 ± 7.9 | 71.8 ± 7.9 | 71.6 ± 7.8 | 0.829 |
| Female sex, n (%) | 194 (47.0) | 161 (47.4) | 33 (45.2) | 0.739 |
| Body mass index, kg/m<sup>2</sup> | 27.3 ± 5.0 | 26.9 ± 4.7 | 29.5 ± 5.6 | <0.001 |
| **Cohort, n (%)** |  |  |  |  |
| Healthy control | 76 (18.4) | 65 (19.1) | 11 (15.1) | 0.040 |
| Prodromal | 145 (35.1) | 110 (32.4) | 35 (47.9) |  |
| Parkinson's disease | 192 (46.5) | 165 (48.5) | 27 (37.0) |  |
| **Vascular factors, n (%)** |  |  |  |  |
| Hypertension | 151 (36.6) | 109 (32.1) | 42 (57.5) | <0.001 |
| Hyperlipidemia | 183 (44.3) | 138 (40.6) | 45 (61.6) | 0.001 |
| Obstructive sleep apnea | 43 (10.4) | 28 (8.2) | 15 (20.5) | 0.002 |
| Obesity (BMI ≥ 30) | 103 (24.9) | 69 (20.3) | 34 (46.6) | <0.001 |
| Stroke/TIA | 16 (3.9) | 15 (4.4) | 1 (1.4) | 0.222 |
| Coronary artery disease | 28 (6.8) | 23 (6.8) | 5 (6.8) | 0.979 |
| Atrial fibrillation | 19 (4.6) | 17 (5.0) | 2 (2.7) | 0.403 |
| **Vascular Burden Score (0–7)** | 1.31 ± 1.14 | 1.17 ± 1.10 | 1.97 ± 1.13 | <0.001 |
| Microvascular (0–4) | 1.16 ± 1.03 | 1.01 ± 0.96 | 1.86 ± 1.08 | <0.001 |
| Macrovascular (0–3) | 0.15 ± 0.38 | 0.16 ± 0.39 | 0.11 ± 0.32 | 0.291 |

BMI = body mass index; DM = diabetes mellitus; Pre-DM = prediabetes; SD = standard deviation; TIA = transient ischemic attack.

Modified Poisson regression with robust variance, adjusted for age, sex, and body mass index, identified three microvascular factors surviving Benjamini-Hochberg false discovery rate correction (Table 2). As seen in Figure 2, obesity showed the strongest amplification (aPR 2.28, 95% confidence interval 1.66 to 3.15, p < 0.001, q = 0.000041). Hypertension was elevated (aPR 1.60, 95% confidence interval 1.25 to 2.05, p < 0.001, q = 0.002331), as was hyperlipidemia (aPR 1.45, 95% confidence interval 1.16 to 1.83, p = 0.001, q = 0.005705). Obstructive sleep apnea showed elevation that did not survive false discovery rate correction (aPR 1.52, 95% confidence interval 0.80 to 2.89, p = 0.205). Macrovascular factors showed no significant association: stroke or transient ischemic attack (aPR 0.39, 95% confidence interval 0.05 to 2.80, p = 0.346), coronary artery disease (aPR 0.91, 95% confidence interval 0.36 to 2.29, p = 0.834), and atrial fibrillation (aPR 0.42, 95% confidence interval 0.10 to 1.79, p = 0.240).

**Table 2.** Per-factor adjusted prevalence ratios for diabetes mellitus/prediabetes association with vascular risk factors (N = 413). Modified Poisson regression with robust variance, adjusted for age, sex, and body mass index. Benjamini-Hochberg false discovery rate correction applied across the seven factors. Bolded factors survived false discovery rate correction at q < 0.05.

| Vascular factor | Category | aPR (95% CI) | p-value | q-value (BH-FDR) |
| --- | --- | --- | --- | --- |
| **Obesity (BMI $\geq 30$)** | Microvascular | **2.28 (1.66–3.15)** | **<0.001** | **<0.001** |
| **Hypertension** | Microvascular | **1.60 (1.25–2.05)** | **<0.001** | **0.002** |
| **Hyperlipidemia** | Microvascular | **1.45 (1.16–1.83)** | **0.001** | **0.006** |
| Sleep apnea (OSA) | Microvascular | 1.52 (0.80–2.89) | 0.205 | 0.287 |
| Stroke/TIA | Macrovascular | 0.39 (0.05–2.80) | 0.346 | 0.404 |
| Coronary artery disease | Macrovascular | 0.91 (0.36–2.29) | 0.834 | 0.834 |
| Atrial fibrillation | Macrovascular | 0.42 (0.10–1.79) | 0.240 | 0.287 |

aPR = adjusted prevalence ratio; CI = confidence interval; BH-FDR = Benjamini-Hochberg false discovery rate; BMI = body mass index; OSA = obstructive sleep apnea; TIA = transient ischemic attack. Reference group: participants without diabetes mellitus or prediabetes ( $n = 340$ ). Wide confidence intervals for stroke/TIA, coronary artery disease, and atrial fibrillation reflect small event counts in the diabetes mellitus/prediabetes group (1, 5, and 2 events respectively).

**Figure 2.**
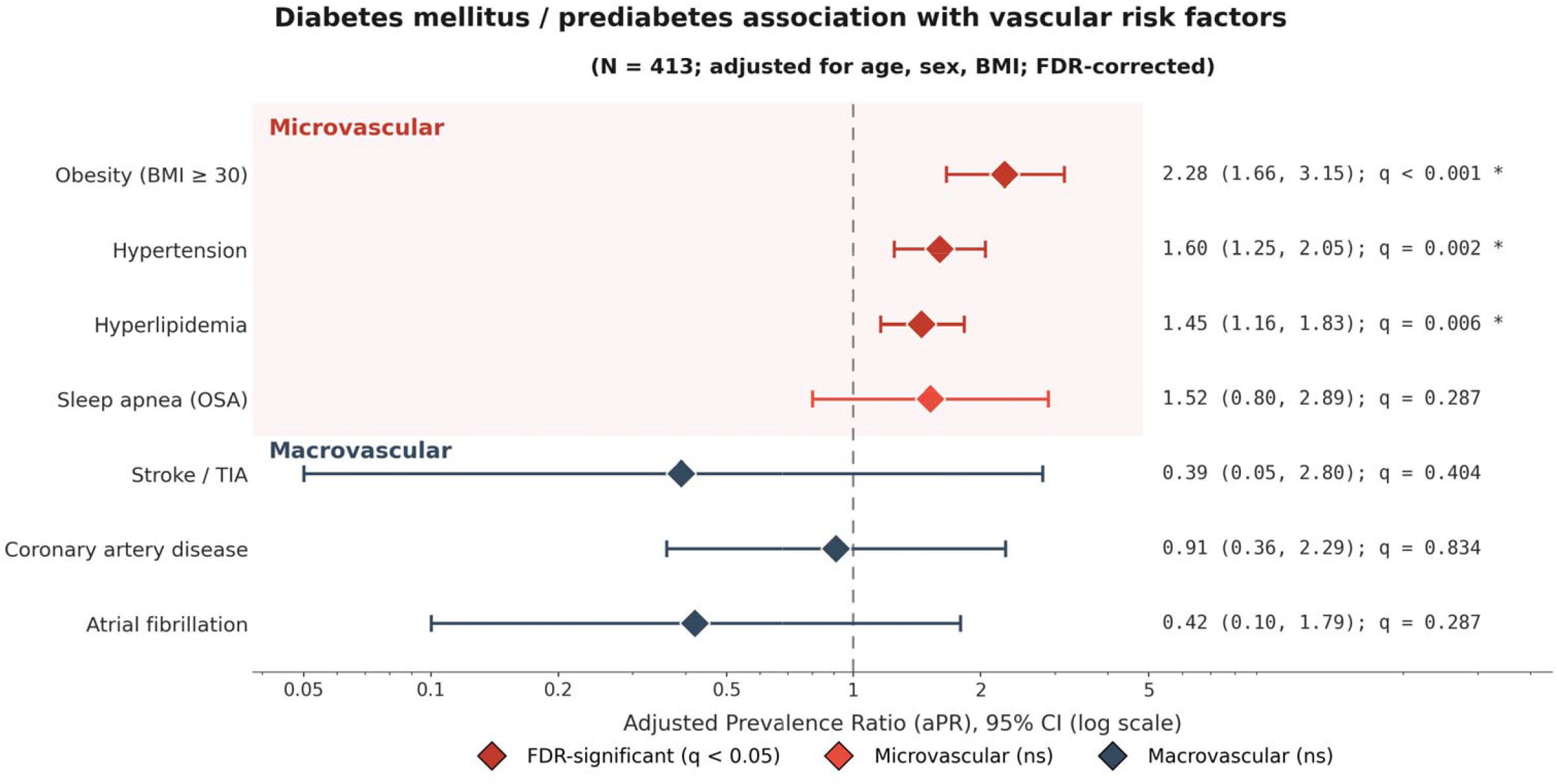
Per-Factor Adjusted Prevalence Ratios for Diabetes Mellitus / Prediabetes Association with Vascular Risk Factors. Forest plot of age-, sex-, and body mass index-adjusted prevalence ratios (aPR) for each of seven vascular risk factors comparing diabetes mellitus/prediabetes participants (n = 73) versus participants without diabetes mellitus/prediabetes (n = 340) in the PPMI sample (N = 413). For obesity, body mass index was omitted from the covariate set to avoid collinearity. Modified Poisson regression with robust variance was used to estimate aPRs, with Benjamini-Hochberg false discovery rate correction applied across the seven factors. Three microvascular factors survived FDR correction (q < 0.05, marked with asterisks): obesity (aPR 2.28), hypertension (aPR 1.60), and hyperlipidemia (aPR 1.45). Sleep apnea attenuated to non-significance after body mass index adjustment, consistent with shared adiposity confounding. Macrovascular factors (stroke/TIA, coronary artery disease, atrial fibrillation) showed no diabetic amplification, with wide confidence intervals reflecting small event counts. The dashed vertical line indicates aPR = 1 (no association); prevalence ratios presented on log scale.

Multivariable linear regressions of total Vascular Burden Score and the microvascular and macrovascular sub-scores (Models A, B, C; **see Supplementary Table S2**) examined the composite phenotype. The macrovascular sub-score regression was the analytic anchor for testing whether diabetic amplification reflects metabolic syndrome overlapping pathology or extends to end-organ atherothrombotic events. After adjustment for age, sex, and body mass index, the diabetes mellitus/prediabetes effect on the macrovascular sub-score was non-significant (β = −0.06, 95% confidence interval −0.15 to 0.04, p = 0.251), in contrast to the strong amplification observed for the microvascular sub-score (β = 0.85, 95% confidence interval 0.60 to 1.09, p < 0.001)(Figure 3B). The total Vascular Burden Score regression confirmed the concentration of microvascular contribution with simultaneous diminished macrovascular contribution (β = 0.53, 95% confidence interval 0.29 to 0.77, p < 0.001, model R^2^ = 0.338). Because macrovascular events were rare, this null finding should be interpreted as absence of detectable enrichment rather than definitive evidence of macrovascular sparing. Full regression results are presented in Supplementary Table S2. Count-model sensitivity analyses (Supplementary Table S5) yielded substantively concordant inference across all three models, supporting the validity of the linear-model specification.

A two-way analysis of variance examining Vascular Burden Score as a function of cohort by diabetes mellitus/prediabetes, adjusted for age, sex, and body mass index, revealed a significant diabetes mellitus main effect (F(1, 404) = 17.98, p < 0.001, partial η^2^= 0.043) and a non-significant cohort by diabetes mellitus interaction (F(2, 404) = 0.29, p = 0.747, partial η^2^= 0.001). Cohort itself did not independently predict Vascular Burden Score after covariate adjustment (F(2, 404) = 0.86, p = 0.426). Within-cohort comparisons confirmed comparable diabetic effect magnitudes in all conditions: healthy controls Cohen’s d = 1.08 (p = 0.009), prodromal d = 0.58 (p = 0.005), and PD d = 0.75 (p = 0.001) (Figure 3A). The non-significant interaction indicates no detectable evidence that the diabetes/prediabetes-associated VBS elevation differed across PPMI diagnostic cohorts. This pattern is consistent with the possibility that cardiometabolic vascular-risk burden is present before clinical PD diagnosis, but longitudinal follow-up is required to establish temporal precedence.

**Figure 3.**
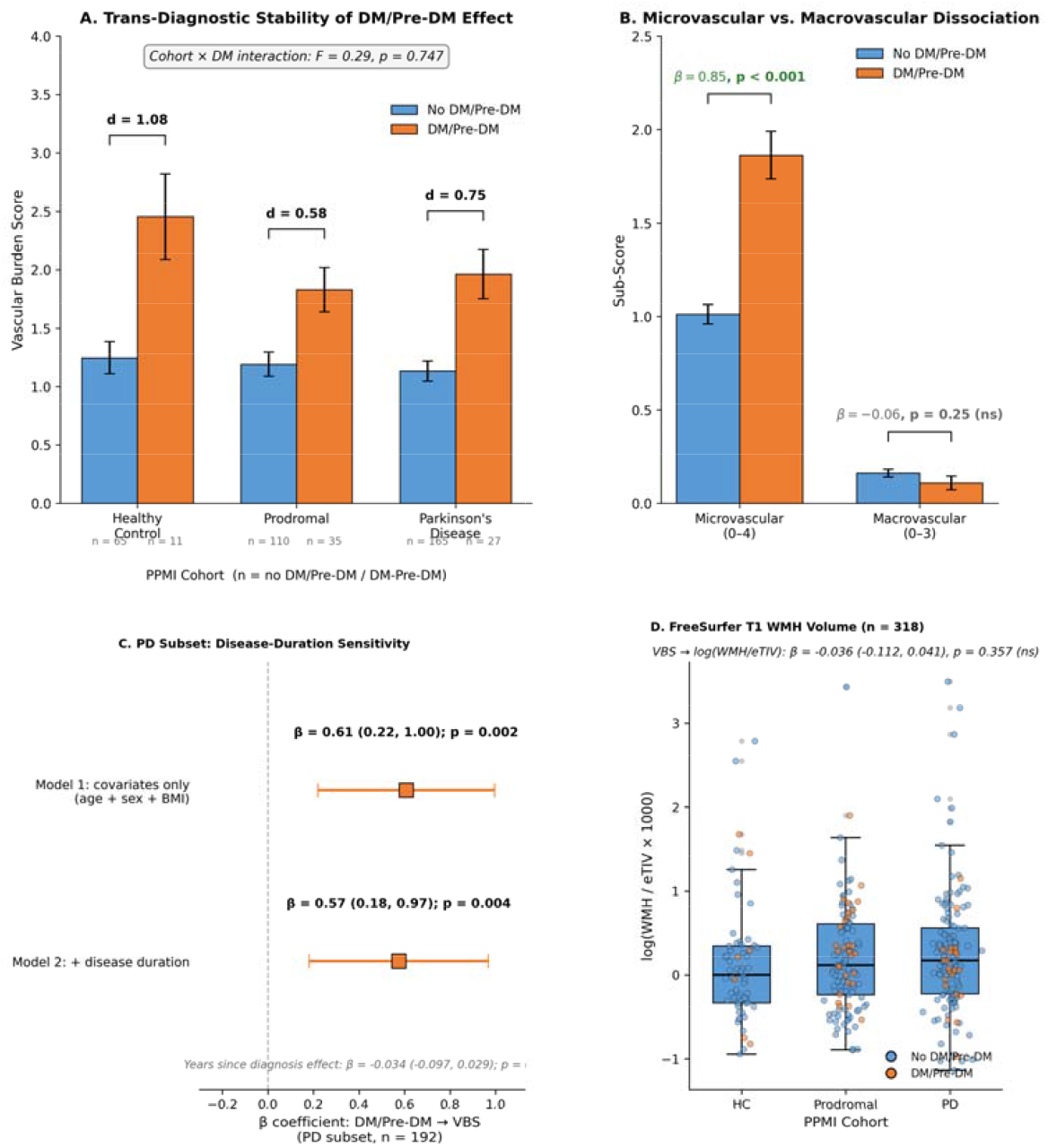
Across Diagnostic Stage Stability and Sensitivity Analyses of the Diabetes-Associated Vascular Burden Phenotype. (A) Vascular Burden Score by diabetes mellitus/prediabetes status, stratified by PPMI clinical cohort (healthy control, prodromal, Parkinson’s disease). Within-cohort Cohen’s d effect sizes (1.08, 0.58, 0.75) demonstrate comparable diabetic amplification across the diagnostic spectrum; the cohort by diabetes mellitus interaction was non-significant (F = 0.29, p = 0.747). (B) Microvascular sub-score and macrovascular sub-score by diabetes mellitus/prediabetes status across the full sample. The diabetes mellitus/prediabetes effect concentrates in the microvascular sub-score (β = 0.85, p < 0.001) and is absent for the macrovascular sub-score (β = −0.06, p = 0.25), establishing the microvascular versus macrovascular dissociation that is the analysis’s primary non-circular finding. Error bars represent standard error of the mean. (C) Disease duration sensitivity: forest plot of standardized regression coefficients for the diabetes mellitus/prediabetes effect on Vascular Burden Score in the PD subset (n = 192) with and without disease duration as a covariate, demonstrating that the diabetes mellitus/prediabetes association is independent of years since diagnosis. (D) Box plot of FreeSurfer 7 derived white matter hypointensity volume (normalized to estimated total intracranial volume, log scale) by clinical cohort in the 318 participants with FreeSurfer data available. Distributions overlap across cohorts; the regression of log-transformed normalized white matter hypointensity volume on Vascular Burden Score showed no significant association after adjustment for age, sex, and body mass index (β = −0.036, p = 0.357). AUC = area under the receiver operating characteristic curve; DTI = diffusion tensor imaging; FS = FreeSurfer; SN = substantia nigra; VBS = Vascular Burden Score; WMH = white matter hypointensities.

The first sensitivity analysis testing diabetic/prediabetic effect after adjusting for date of PD diagnosis showed the diabetes mellitus/prediabetes effect remained significant (β = 0.574, p = 0.0044) and disease duration was not associated with Vascular Burden Score (β = -0.034, p = 0.291), indicating that the observed phenotype is not driven by PD disease duration (Figure 3C).

A second sensitivity analysis examined whether Vascular Burden Score is associated with structural cerebrovascular damage measurable on T1 imaging. Among 318 participants with FreeSurfer 7 ASEG output available, the total Vascular Burden Score was not significantly associated with log-transformed normalized white matter hypointensity volume after adjustment for age, sex, and body mass index (β = -0.036, 95% confidence interval −0.112 to 0.041, p = 0.357; corresponding to −3.5% change per Vascular Burden Score point, 95% confidence interval −10.6% to +4.1%). Age strongly predicted white matter hypointensity volume (p < 0.001). The null association is interpreted in the context of T1 hypointensity sensitivity limitations and PPMI cerebrovascular burden floor effects (Figure 3D; Supplementary Analysis S1).

## Discussion

This analysis had three aims: to characterize the diabetes mellitus and prediabetes associated vascular phenotype using a composite Vascular Burden Score, test whether the phenotype operates through microvascular versus macrovascular pathways, and evaluate whether the phenotype is stable across PPMI diagnostic cohorts. The Vascular Burden Score successfully captured cardiometabolic vascular-risk phenotype associated with diabetes mellitus and prediabetes in PPMI. The composite phenotype emerged across three converging findings: a microvascular elevation surviving false discovery rate correction (obesity aPR 2.28, hypertension aPR 1.60, hyperlipidemia aPR 1.45), diminished macrovascular contribution across stroke, coronary artery disease, and atrial fibrillation (β = −0.06, p = 0.25), and statistically unchanged within-cohort effect sizes spanning healthy controls (Cohen’s d = 1.08), prodromal individuals (d = 0.58), and clinically diagnosed PD (d = 0.75). The microvascular clustering between type 2 diabetes mellitus and obesity, hypertension, and hyperlipidemia are interrelated by metabolic syndrome definitional criteria.^20,22^ The non-circular contribution of this analysis is the absence of a macrovascular signal: diabetic amplification has not propagated to the three clinically manifest macrovascular conditions assessed here (stroke/TIA, coronary artery disease, and atrial fibrillation) at this cohort stage.

Non-significant interactions are typically treated as methodological controls supporting pooled analysis, but in this context the absence of a cohort by diabetes mellitus/prediabetes interaction (F = 0.29, p = 0.747) suggests that the association between diabetes/prediabetes and VBS did not detectably differ among controls, prodromal participants, and clinically diagnosed PD participants. Comparable within-cohort effect sizes are consistent with a cardiometabolic vascular-risk phenotype that is not restricted to established PD. However, because the analysis is cross-sectional, longitudinal data are needed to determine whether this phenotype precedes PD onset or predicts progression. This across diagnostic stage stability extends prior PD vascular characterization into the prodromal stage, where Movement Disorder Society research criteria define a population at elevated PD risk through hyposmia, REM sleep behavior disorder, and genetic variants before motor symptom onset,^18^ positioning composite cardiometabolic phenotyping as a candidate antecedent risk-stratification dimension complementing existing prodromal markers, extending health-record-derived risk-algorithm approaches that have been applied to PD prediction in primary care settings.^37^ Notably, the prodromal participants in our analytic sample were enrolled exclusively as non-manifesting carriers of pathogenic LRRK2 (n = 91) or GBA (n = 60) variants.^32^ Building on the foundational VADO study, which first identified diabetes mellitus as the singular vascular risk factor differentially associated with vascular versus idiopathic parkinsonism,^24^ the present analysis extends that observation to a composite phenotyping framework demonstrable across PPMI diagnostic cohorts.

The microvascular concentration and macrovascular sparing observed in PPMI may be consistent with a framework in which chronic hyperglycemia contributes to progressive endothelial dysfunction, blood-brain barrier compromise, and small vessel pathology before clinically apparent atherothrombotic events emerge, a pattern that could potentially involve renin-angiotensin system mediated oxidative stress.^8,26,27^ Mendelian randomization evidence has established that type 2 diabetes mellitus exerts a causal effect on cerebral small vessel disease, with strongest signals on lacunar stroke and white matter integrity,^8^ providing support for the microvascular pathway our composite phenotyping framework operationalizes. Exploratory random forest classification testing Vascular Burden Score components against the three derived nigral diffusion metrics (FA, MD, RD) yielded modest performance in the small imaging subset (full model AUC = 0.515; Supplementary Analysis S2), with permutation importance dominated by nigral diffusion features and smaller but non-zero contributions from Vascular Burden Score sub-scores, consistent with capture of variance not redundant with nigral microstructure.

The Vascular Burden Score offers a reproducible quantitative tool computable from routine clinical data, requiring only medical history flags and body mass index without specialty imaging or advanced laboratory assays building on composite vascular risk scoring approaches established in cognitive aging research.^36^ Because the score is based on clinical risk-factor indicators rather than direct vascular imaging, it should be interpreted as a vascular-risk phenotyping tool rather than a direct measure of cerebral small vessel disease burden. For prodromal PD populations identified via Movement Disorder Society research criteria,^18,25^ VBS adds a vascular phenotyping dimension complementing existing motor and non-motor prodromal markers and supports integrated multimodal risk stratification at the disease stage where intervention has greatest potential to modify trajectory before clinical motor symptom onset. The across diagnostic stage stability finding motivates investigation of vascular phenotyping in prodromal and at-risk populations, where contemporary intervention frameworks have proposed multicomponent management of cardiovascular risk factors as candidate disease-modifying therapy.^6^

Strengths of this analysis include a multi-site PD biomarker cohort with standardized phenotyping, modified Poisson regression appropriate to a common cross-sectional outcome, the pre-specified microvascular and macrovascular partition, and complementary nigral diffusion and FreeSurfer-derived sensitivity analyses.^21,30^ Several limitations warrant acknowledgment. The cross-sectional design precludes definitive causal inference about temporal precedence; observed cohort invariance is consistent with antecedent interpretation but cannot establish it within individuals. Construct overlap between metabolic syndrome components and type 2 diabetes mellitus motivated the microvascular and macrovascular partition structure but cannot be eliminated entirely.^20,22,23^ Accordingly, the association between diabetes/prediabetes and the microvascular sub-score should be interpreted partly as confirmation of expected cardiometabolic clustering rather than discovery of a novel diabetes-specific vascular syndrome. Ascertainment via the PPMI Medical Conditions Log relies on clinician-entered coding subject to detection bias, though this would not affect the diminished macrovascular contribution.^17^ PPMI participants are predominantly non-Hispanic White and well-educated relative to the broader PD population, limiting generalizability. Additionally, the PPMI prodromal cohort in our analytic sample comprised exclusively non-manifesting genetic risk variant carriers (predominantly LRRK2 and GBA); generalizability of our findings to idiopathic prodromal PD populations defined by REM sleep behavior disorder or hyposmia remains to be established.^17^ Our macrovascular sub-score captures three cardiovascular conditions (stroke/TIA, coronary artery disease, atrial fibrillation) but does not include other clinically important end-organ manifestations of diabetes mellitus such as chronic kidney disease, diabetic retinopathy, peripheral artery disease, or diabetic neuropathy. The exploratory white matter hypointensity analysis did not show significant association with Vascular Burden Score, which may reflect the limited sensitivity of T1 based detection for early small vessel disease and the relatively low cerebrovascular burden in PPMI (median normalized white matter hypointensity volume 1.12 mL per L estimated total intracranial volume); replication using T2 fluid-attenuated inversion recovery white matter hyperintensity quantification in cohorts with greater cerebrovascular burden is warranted.^5,7^ The supplementary random forest analysis was limited to controls and PD participants with diffusion tensor imaging (n = 50); prodromal participants did not have baseline diffusion tensor imaging in the available PPMI release, precluding three-class classification. Macrovascular events were rare (atrial fibrillation prevalence below 5% in non-diabetic participants), limiting power for detecting modest diabetic effects on these outcomes. Therefore, no significant macrovascular findings should be interpreted cautiously and do not exclude modest associations in larger or higher-risk cohorts. Future longitudinal characterization of Vascular Burden Score trajectories within individuals followed from healthy or prodromal status through clinical PD onset would directly address temporal precedence, extending prior cohort evidence linking diabetes mellitus to incident PD risk.^38^ External validations in cohorts with greater cardiometabolic burden and demographic diversity than PPMI would address generalizability.

In conclusion, among 413 PPMI participants, diabetes mellitus and prediabetes were associated with elevated composite microvascular burden, no parallel macrovascular contribution, and comparable effect magnitudes across healthy controls, prodromal individuals, and clinically diagnosed PD. The Vascular Burden Score offers a reproducible clinical phenotyping tool that may complement established disease-specific biomarkers in early-stage parkinsonian risk stratification, pending longitudinal validation.

## Author Contributions

**A.B**.: 1A, 1B, 1C, 2A, 2B, 3A.

**S.Y.C**.: 1A, 1B, 2A, 3B.

**K.C**.: 1A, 2A, 2C, 3B.

**R.T**.: 1A, 1B, 3B.

**E.O**.: 1A, 1B, 2A, 2C, 3B (supervision and corresponding author).

*Author contribution categories per Movement Disorders convention: (1) Research Project: A. Conception, B. Organization, C. Execution; (2) Statistical Analysis: A. Design, B. Execution, C. Review and Critique; (3) Manuscript: A. Writing of the first draft, B. Review and Critique*.

## Financial Disclosures (Past 12 Months)

**A. Belnavis:** Receives stipend support from Arizona State University College of Health Solutions. No commercial, governmental, or foundation financial relationships related to this work.

**S.Y. Chiu:** Salaried employment at Mayo Clinic. No commercial financial relationships related to this work.

**K. Chen:** Salaried employment at Arizona State University. No commercial financial relationships related to this work.

**R. Thorpe:** Salaried employment at Johns Hopkins University. Dr. Thorpe was supported by NIA P30AG059298. No commercial financial relationships related to this work.

**E. Ofori:** Salaried employment at Arizona State University. Research support from internal ASU College of Health Solutions funds. No commercial financial relationships related to this work.

## Funding

This research received support from Arizona State University College of Health Solutions. Data were accessed through the PPMI Data Use Agreement. The funders had no role in study design, data analysis, interpretation, or manuscript preparation.

### Ethical Compliance Statement

This analysis of de-identified PPMI data received institutional review board exemption from Arizona State University. The original PPMI study received institutional review board approval at all participating sites and informed consent was obtained from all subjects. The authors confirm we have read the Journal’s position on issues involved in ethical publication and affirm that this work is consistent with those guidelines.

## Data Availability Statement

Data used in the preparation of this article were obtained on February 12, 2026 from the Parkinson’s Precision Medicine Initiative (PPMI) database (www.ppmi-info.org/access-data-specimens/download-data), RRID:SCR_006431. For up-to-date information on the study, visit www.ppmi-info.org. This analysis used data openly available from PPMI (Tier 1 Data). Analysis code is deposited at https://github.com/asbelnav-source/VBS_MDS_05-2026 and will be made publicly accessible upon publication.

## Acknowledgments

PPMI – a public-private partnership – is funded by the Michael J. Fox Foundation for Parkinson’s Research and funding partners, including 4D Pharma, AbbVie, AcureX, Allergan, Amathus Therapeutics, Aligning Science Across Parkinson’s, AskBio, Avid Radiopharmaceuticals, BIAL, BioArctic, Biogen, Biohaven, BioLegend, BlueRock Therapeutics, Bristol-Myers Squibb, Calico Labs, Capsida Biotherapeutics, Celgene, Cerevel Therapeutics, Coave Therapeutics, DaCapo Brainscience, Denali, Edmond J. Safra Foundation, Eli Lilly, Gain Therapeutics, GE HealthCare, Genentech, GSK, Golub Capital, Handl Therapeutics, Insitro, Jazz Pharmaceuticals, Johnson & Johnson Innovative Medicine, Lundbeck, Merck, Meso Scale Discovery, Mission Therapeutics, Neurocrine Biosciences, Neuron23, Neuropore, Pfizer, Piramal, Prevail Therapeutics, Roche, Sanofi, Servier, Sun Pharma Advanced Research Company, Takeda, Teva, UCB, Vanqua Bio, Verily, Voyager Therapeutics, the Weston Family Foundation and Yumanity Therapeutics. The authors thank the PPMI study participants and the PPMI study sponsors and investigators.

