## Supplemental Table 1 for "Vascular Phenotyping in Parkinson’s Disease: Diabetes Mellitus Operationalizes a Microvascular Metabolic Syndrome Cluster Across PPMI Diagnostic Cohorts"

**Contents:**

Supplementary Table S1. Baseline clinical characteristics by cohort and diabetes mellitus/prediabetes status.

Supplementary Table S2. Multivariable linear regression of composite Vascular Burden Scores.

Supplementary Table S3. Across diagnostic stage stability of diabetes mellitus/prediabetes effect on Vascular Burden Score (ANOVA decomposition and within-cohort effect sizes).

Supplementary Table S4. Sensitivity analysis: PD-only subset with disease duration.

Supplementary Table S5 Count-model sensitivity analyses for composite Vascular Burden Score outcomes.

Supplementary Table S6 Random forest classification performance and permutation variable importance for the exploratory analysis evaluating Vascular Burden Score components against nigral diffusion metrics.

Supplementary Analysis S1. FreeSurfer T1 white matter hypointensity volume analysis.

Supplementary Analysis S2. Random Forest Sensitivity Analysis Against Nigral Diffusion Metrics.

Supplementary Figure S1. Vascular factor prevalence by diabetes mellitus/prediabetes status across the PPMI clinical spectrum.

Supplementary Figure S2. Random Forest Permutation Variable Importance.

### Supplementary Table S1

**Baseline clinical characteristics by cohort and diabetes mellitus/prediabetes status (N = 413).**

Detailed clinical phenotyping of the 413-participant analytic sample, stratified by PPMI clinical cohort (healthy control, prodromal, clinically diagnosed Parkinson's disease) and diabetes mellitus/prediabetes status. The PPMI prodromal cohort in this sample consisted entirely of non-manifesting carriers of pathogenic variants in LRRK2 or GBA who met PPMI's genetic-prodromal enrollment criteria; participants enrolled via idiopathic REM sleep behavior disorder or hyposmia pathways were not represented. Disease-severity measures (MDS-UPDRS, Hoehn & Yahr) are interpreted in the context of an early-stage clinical PD cohort under standard medication conditions. Continuous variables presented as mean ± SD; categorical variables as n (%); Hoehn & Yahr stage as median (interquartile range).

| **Characteristic** | **Healthy Control** | **Healthy Control** | **Prodromal** | **Prodromal** | **PD** | **PD** |
| --- | --- | --- | --- | --- | --- | --- |
|  | **No DM/Pre-DM (n = 65)** | **DM/Pre-DM (n = 11)** | **No DM/Pre-DM (n = 110)** | **DM/Pre-DM (n = 35)** | **No DM/Pre-DM (n = 165)** | **DM/Pre-DM (n = 27)** |
| ***Demographics*** |  |  |  |  |  |  |
| Age, years | 74.7 ± 9.3 | 76.4 ± 7.9 | 70.6 ± 7.1 | 70.5 ± 7.1 | 71.5 ± 7.7 | 71.0 ± 8.2 |
| Female sex, n (%) | 32 (49.2%) | 2 (18.2%) | 53 (48.2%) | 20 (57.1%) | 76 (46.1%) | 11 (40.7%) |
| Body mass index, kg/m² | 25.9 ± 4.3 | 30.1 ± 3.8 | 27.1 ± 5.3 | 29.6 ± 6.1 | 27.1 ± 4.4 | 29.1 ± 5.8 |
| White race, n (%) | 63 (96.9%) | 11 (100.0%) | 109 (99.1%) | 35 (100.0%) | 163 (98.8%) | 25 (92.6%) |
| ***Disease severity*** |  |  |  |  |  |  |
| Years since PD diagnosis | N/A | N/A | N/A | N/A | 11.6 ± 2.1 | 10.7 ± 2.4 |
| MDS-UPDRS Part I | 0.6 ± 1.3 | 0.5 ± 0.9 | 1.0 ± 1.4 | 1.8 ± 2.1 | 1.5 ± 1.9 | 1.6 ± 1.7 |
| MDS-UPDRS Part II | 0.5 ± 0.9 | 0.5 ± 1.5 | 0.8 ± 1.7 | 1.5 ± 2.8 | 6.3 ± 4.3 | 5.8 ± 4.8 |
| MDS-UPDRS Part III (motor) | 1.4 ± 2.3 | 1.0 ± 1.1 | 2.3 ± 3.1 | 3.2 ± 3.9 | 20.6 ± 9.8 | 24.4 ± 12.9 |
| Hoehn & Yahr stage | 0.0 (0.0, 0.0) | 0.0 (0.0, 0.0) | 0.0 (0.0, 0.0) | 0.0 (0.0, 0.0) | 2.0 (1.0, 2.0) | 2.0 (1.0, 2.0) |
| MoCA total | 28.2 ± 1.1 | 28.2 ± 1.0 | 26.8 ± 2.3 | 26.4 ± 2.4 | 26.6 ± 2.5 | 26.1 ± 2.8 |
| ***Genetic variant carrier status*** |  |  |  |  |  |  |
| LRRK2, n (%) | 0 (0.0%) | 0 (0.0%) | 69 (62.7%) | 22 (62.9%) | 70 (42.4%) | 14 (51.9%) |
| GBA, n (%) | 0 (0.0%) | 0 (0.0%) | 46 (41.8%) | 14 (40.0%) | 42 (25.5%) | 6 (22.2%) |
| Other (SNCA/PRKN/VPS35/PINK1), n (%) | 0 (0.0%) | 0 (0.0%) | 0 (0.0%) | 0 (0.0%) | 1 (0.6%) | 0 (0.0%) |
| Any PD-associated variant, n (%) | 0 (0.0%) | 0 (0.0%) | 110 (100.0%) | 35 (100.0%) | 110 (66.7%) | 20 (74.1%) |
| ***Prodromal phenotyping (descriptive)*** |  |  |  |  |  |  |
| RBDSQ total score | 2.1 ± 1.8 | 3.0 ± 2.6 | 2.6 ± 2.0 | 2.5 ± 2.3 | 3.3 ± 2.8 | 2.6 ± 2.6 |
| UPSIT total score | — | — | 32.7 ± 3.2 | 32.0 (n=1) | — | — |
| ***Vascular phenotype*** |  |  |  |  |  |  |
| Vascular Burden Score (0–7) | 1.25 ± 1.10 | 2.45 ± 1.21 | 1.19 ± 1.08 | 1.83 ± 1.12 | 1.13 ± 1.11 | 1.96 ± 1.09 |
| Microvascular sub-score (0–4) | 1.02 ± 0.94 | 2.09 ± 1.04 | 1.09 ± 1.00 | 1.74 ± 1.09 | 0.96 ± 0.93 | 1.93 ± 1.11 |
| Macrovascular sub-score (0–3) | 0.23 ± 0.46 | 0.36 ± 0.50 | 0.10 ± 0.30 | 0.09 ± 0.28 | 0.18 ± 0.41 | 0.04 ± 0.19 |

*Continuous variables presented as mean ± SD unless otherwise noted; categorical variables as n (%); Hoehn & Yahr stage as median (interquartile range). Body mass index ≥ 30 kg/m² defines the obesity microvascular factor and is the dominant driver of within-cohort BMI differences between diabetes mellitus/prediabetes groups. RBDSQ = REM Sleep Behavior Disorder Screening Questionnaire (range 0–13). UPSIT = University of Pennsylvania Smell Identification Test (number of items correct out of 40); UPSIT scores are presented for the prodromal cohort only because UPSIT testing in PPMI is concentrated at the screening visit, with limited UPSIT data available for healthy controls and clinically diagnosed PD participants at baseline and with only one prodromal diabetes mellitus/prediabetes participant having a baseline UPSIT score. MDS-UPDRS Part III scores in clinically diagnosed PD reflect standard medication conditions; on/off-medication scoring was not uniformly available across all participants. The PPMI prodromal cohort in this analytic sample comprised exclusively non-manifesting carriers of pathogenic variants in LRRK2 (n = 91) and/or GBA (n = 60); generalizability to idiopathic REM sleep behavior disorder or hyposmia-defined prodromal populations is not established. BMI = body mass index; DM = diabetes mellitus; MoCA = Montreal Cognitive Assessment; MDS-UPDRS = Movement Disorder Society Unified Parkinson's Disease Rating Scale; PD = Parkinson's disease; Pre-DM = prediabetes; RBDSQ = REM Sleep Behavior Disorder Screening Questionnaire; SD = standard deviation; UPSIT = University of Pennsylvania Smell Identification Test.*

### Supplementary Table S2

**Multivariable linear regression of composite Vascular Burden Scores (N = 413).**

Three linear regression models examined the independent effect of diabetes mellitus/prediabetes on (A) total Vascular Burden Score, (B) microvascular sub-score (hypertension + hyperlipidemia + sleep apnea + obesity), and (C) macrovascular sub-score (stroke/TIA + coronary artery disease + atrial fibrillation). Body mass index omitted from Model B because obesity (BMI ≥ 30) is a component of the microvascular sub-score.

| **Predictor** | **B** | **SE** | **95% CI** | **p** | **R²** |
| --- | --- | --- | --- | --- | --- |
| **Model A: Total VBS (0–7)** |  |  |  |  | 0.338 |
| DM/Pre-DM status | +0.529 | 0.123 | 0.287 to 0.771 | <0.001 |  |
| Age (years) | +0.025 | 0.006 | 0.014 to 0.037 | <0.001 |  |
| Sex (male) | +0.432 | 0.093 | 0.249 to 0.614 | <0.001 |  |
| BMI (kg/m²) | +0.103 | 0.010 | 0.084 to 0.122 | <0.001 |  |
| **Model B: Microvascular (0–4)** |  |  |  |  | 0.157 |
| DM/Pre-DM status | +0.845 | 0.123 | 0.604 to 1.086 | <0.001 |  |
| Age (years) | +0.013 | 0.006 | 0.001 to 0.025 | 0.028 |  |
| Sex (male) | +0.445 | 0.094 | 0.261 to 0.630 | <0.001 |  |
| **Model C: Macrovascular (0–3)** |  |  |  |  | 0.043 |
| DM/Pre-DM status | −0.057 | 0.049 | −0.153 to 0.040 | 0.251 |  |
| Age (years) | +0.008 | 0.002 | 0.003 to 0.012 | 0.001 |  |
| Sex (male) | +0.089 | 0.037 | 0.016 to 0.162 | 0.017 |  |
| BMI (kg/m²) | +0.002 | 0.004 | −0.006 to 0.009 | 0.676 |  |

*B = unstandardized regression coefficient; BMI = body mass index; CI = confidence interval; DM = diabetes mellitus; Pre-DM = prediabetes; SE = standard error; VBS = Vascular Burden Score.*

### Supplementary Table S3

**Across diagnostic stage stability of diabetes mellitus/prediabetes effect on Vascular Burden Score (N = 413).**

Univariate analysis of variance tested whether the diabetes mellitus/prediabetes effect on Vascular Burden Score differs across PPMI clinical groups (healthy control, prodromal, clinically diagnosed PD), adjusting for age, sex, and body mass index. Within-cohort effect sizes computed from independent-samples t-tests with Hedges' small-sample correction.

**Panel A. Two-way ANOVA: VBS ~ Cohort × DM_Group + age + sex + BMI**

| **Source** | **F** | **df** | **p** | **Partial η²** |
| --- | --- | --- | --- | --- |
| Cohort (main effect) | 0.86 | 2, 404 | 0.426 | 0.004 |
| DM_Group (main effect) | 17.98 | 1, 404 | <0.001 | 0.043 |
| **Cohort × DM_Group** | **0.29** | 2, 404 | **0.747** | 0.001 |

**Panel B. Within-cohort effect of diabetes mellitus/prediabetes on Vascular Burden Score**

| **Cohort** | **No DM/Pre-DM** | **DM/Pre-DM** | **Cohen's d** | **Hedges' g** | **p** |
| --- | --- | --- | --- | --- | --- |
| Healthy control | 1.25 ± 1.10 (n=65) | 2.45 ± 1.21 (n=11) | 1.08 | 1.07 | 0.009 |
| Prodromal | 1.19 ± 1.08 (n=110) | 1.83 ± 1.12 (n=35) | 0.58 | 0.58 | 0.005 |
| Parkinson's disease | 1.13 ± 1.11 (n=165) | 1.96 ± 1.09 (n=27) | 0.75 | 0.74 | 0.001 |

*Values for No DM/Pre-DM and DM/Pre-DM columns presented as mean ± standard deviation. Cohen's d effect sizes calculated as (Mean_DM − Mean_NoDM) / SD_pooled, with Hedges' g providing small-sample correction.*

### Supplementary Table S4

**Sensitivity analysis: PD-only subset with disease duration (n = 192).**

Linear regression restricted to PD participants with available disease duration tested whether the diabetes mellitus/prediabetes effect persists after additional adjustment for years since diagnosis. Disease duration anchored to baseline assessment date from PPMI PDDXDT field.

| **Predictor** | **B** | **SE** | **95% CI** | **p** |
| --- | --- | --- | --- | --- |
| DM/Pre-DM status | +0.574 | 0.199 | 0.181 to 0.967 | 0.004 |
| Age (years) | +0.033 | 0.009 | 0.015 to 0.050 | <0.001 |
| Sex (male) | +0.379 | 0.137 | 0.109 to 0.649 | 0.006 |
| BMI (kg/m²) | +0.111 | 0.015 | 0.082 to 0.140 | <0.001 |
| Years since diagnosis | −0.034 | 0.032 | −0.097 to 0.029 | 0.291 |

*Model R² = 0.346. B = unstandardized regression coefficient; BMI = body mass index; CI = confidence interval; DM = diabetes mellitus; Pre-DM = prediabetes; SE = standard error.*

### Supplementary Table S5

**Count-model sensitivity analyses for composite Vascular Burden Score outcomes.**

| **Model** | **Specification** | **β (95% CI)** | **p-value** | **IRR (95% CI)** |
| --- | --- | --- | --- | --- |
| **Model A: Total VBS (0–7)** | Linear (OLS) | +0.529 (+0.288, +0.771) | <0.001 | — |
|  | Poisson (HC0) | +0.296 (+0.160, +0.432) | <0.001 | 1.345 (1.174, 1.541) |
|  | Negative binomial | +0.315 (−0.017, +0.647) | 0.063 | 1.371 (0.983, 1.910) |
| **Model B: Microvascular (0–4)** | Linear (OLS) | +0.845 (+0.604, +1.085) | <0.001 | — |
|  | Poisson (HC0) | +0.602 (+0.447, +0.757) | <0.001 | 1.826 (1.564, 2.131) |
|  | Negative binomial | +0.593 (+0.270, +0.917) | <0.001 | 1.810 (1.309, 2.501) |
| **Model C: Macrovascular (0–3)** | Linear (OLS) | −0.057 (−0.153, +0.040) | 0.251 | — |
|  | Poisson (HC0) | −0.454 (−1.128, +0.220) | 0.187 | 0.635 (0.324, 1.247) |
|  | Negative binomial | −0.478 (−1.300, +0.345) | 0.255 | 0.620 (0.273, 1.412) |

*Multivariable regression coefficients for the diabetes mellitus/prediabetes predictor across three model specifications. Models A and C adjusted for age, sex, and body mass index; Model B (microvascular sub-score) omitted body mass index because obesity (BMI ≥ 30) is a component of the sub-score. Negative binomial fit with α = 1.0. All models n = 413. Pearson dispersion statistics from the Poisson fits were 0.756 (Model A), 0.817 (Model B), and 0.975 (Model C); values below 1.0 indicate no meaningful overdispersion, supporting Poisson as a valid count-model specification. Substantively concordant inference across all three specifications supports the validity of the primary linear-model results. β = unstandardized regression coefficient; CI = confidence interval; HC0 = Huber-White heteroskedasticity-consistent standard errors; IRR = incidence rate ratio (exp(β)); OLS = ordinary least squares; VBS = Vascular Burden Score.*

### Supplementary Table S6

**Random forest classification performance and permutation variable importance for the exploratory analysis evaluating Vascular Burden Score components against nigral diffusion metrics.**

***Classification Performance***

| **Specification** | **n features** | **AUC** | **Balanced accuracy** | **Cohen's kappa** |
| --- | --- | --- | --- | --- |
| Full model (VBS_micro, VBS_macro, FA, MD, RD) | 5 | 0.515 | 0.508 | 0.016 |
| DTI-only (FA, MD, RD) | 3 | 0.571 | 0.549 | 0.098 |
| VBS-only (VBS_micro, VBS_macro) | 2 | 0.282 | 0.360 | −0.274 |

***Permutation Variable Importance (Full Model, mean across 200 iterations)***

| **Feature** | **Mean importance** |
| --- | --- |
| Nigral radial diffusivity (RD) | 0.213 |
| Nigral fractional anisotropy (FA) | 0.144 |
| Nigral mean diffusivity (MD) | 0.119 |
| Vascular Burden Score microvascular sub-score | 0.041 |
| Vascular Burden Score macrovascular sub-score | 0.020 |

*n = 50 (22 healthy controls, 28 clinically diagnosed PD). Random forest classifier with 500 trees, balanced class weights, 5-fold stratified cross-validation, random seed = 42. Permutation variable importance computed with 200 iterations per feature. FA = fractional anisotropy; MD = mean diffusivity, computed as (λ₁ + λ₂ + λ₃) / 3; RD = radial diffusivity, computed as (λ₂ + λ₃) / 2; VBS = Vascular Burden Score.*

### Supplementary Analysis S1

**FreeSurfer T1 white matter hypointensity volume as cerebrovascular substrate surrogate.**

#### Rationale

To probe whether the peripheral cardiometabolic burden captured by Vascular Burden Score is reflected in central structural cerebrovascular pathology, we performed an exploratory analysis using FreeSurfer 7 derived T1 white matter hypointensity volume as a structural surrogate for cerebral small vessel disease burden. T1 white matter hypointensities are validated against T2 FLAIR white matter hyperintensities and have been used as a small vessel disease surrogate in prior PPMI imaging analyses; they are less sensitive than FLAIR for early lesions but more specific for established tissue damage.

#### Methods

Baseline FreeSurfer 7 ASEG output was obtained for 1,713 PPMI participants; 318 of the 413 primary-analysis participants had FreeSurfer-derived white matter hypointensity volume available. Hypointensity volume (mm³) was normalized to estimated total intracranial volume and reported as mL per L estimated total intracranial volume. Log-transformed normalized volumes were used in models to address right-skew. Linear regression of log-transformed normalized white matter hypointensity volume on (a) total Vascular Burden Score and (b) microvascular and macrovascular sub-scores separately, adjusted for age, sex, and body mass index. Stratified analyses by Cohort and DM_Group were also conducted.

#### Results

Cohort distribution among the 318 participants with FreeSurfer data: healthy control n = 61, prodromal n = 118, PD n = 139. Diabetes mellitus/prediabetes n = 59. White matter hypointensity volume showed median 1,741 mm³ (IQR 1,166 to 2,585); normalized median 1.12 mL per L estimated total intracranial volume (IQR 0.78 to 1.69).

Vascular Burden Score was not significantly associated with white matter hypointensity volume after adjustment for age, sex, and body mass index (β = −0.036, 95% confidence interval −0.112 to 0.041, p = 0.357; corresponding to −3.5% per Vascular Burden Score point, 95% confidence interval −10.6% to +4.1%). Component-level analyses showed similar null findings: microvascular sub-score (−2.8% per point, 95% confidence interval −9.6% to +4.5%, p = 0.442) and macrovascular sub-score (−3.2% per point, 95% confidence interval −21.0% to +18.6%, p = 0.756). Age was strongly associated with white matter hypointensity volume (p < 0.001). Diabetes mellitus/prediabetes as a standalone predictor was not associated with white matter hypointensity volume (β = 0.027, p = 0.782, adjusted). Cohort-stratified analyses were null in healthy controls (p = 0.806), prodromal (p = 0.484), and PD (p = 0.200).

#### Interpretation

The null association between Vascular Burden Score and white matter hypointensity volume has three plausible explanations, which are not mutually exclusive. First, PPMI is an enriched cohort with relatively low cerebrovascular disease burden (median normalized white matter hypointensity volume approximately 1.1 mL per L estimated total intracranial volume; population studies of similar age range typically report 3 to 5 times this volume), and floor effects may limit detection of real but small associations. Second, T1 white matter hypointensities reflect established tissue damage and are less sensitive than FLAIR hyperintensities for early or subtle small vessel disease; an association between Vascular Burden Score and earlier-stage cerebrovascular change would be missed by this surrogate. Third, Vascular Burden Score may capture peripheral cardiometabolic risk burden that has not yet, or only partially, translated into measurable structural cerebrovascular damage in this relatively early-stage cohort.

**Supplementary Analysis S2. Random Forest Sensitivity Analysis Against Nigral Diffusion Metrics**

Whether Vascular Burden Score components capture variance distinct from canonical nigral diffusion microstructural biomarkers was tested using a random forest classifier (500 trees, balanced class weights) in the subset of PPMI participants with both baseline diffusion tensor imaging and complete vascular phenotyping (n = 50; 22 healthy controls and 28 clinically diagnosed PD). No prodromal participants had baseline diffusion tensor imaging in the available PPMI release, precluding three-class classification. Five features were entered: Vascular Burden Score microvascular sub-score, Vascular Burden Score macrovascular sub-score, and three nigral diffusion metrics derived from the eigenvalues produced by the PPMI CIND pipeline. The diffusion metrics were fractional anisotropy (FA), mean diffusivity (MD), and radial diffusivity (RD).^29,34^ Model performance was evaluated under 5-fold stratified cross-validation with the area under the receiver operating characteristic curve (AUC), balanced accuracy, and Cohen's kappa as primary metrics. Feature contributions were quantified via permutation variable importance (200 iterations per feature). To isolate the incremental contribution of Vascular Burden Score components, AUCs were also computed for DTI-only (FA, MD, RD) and Vascular Burden Score-only (microvascular and macrovascular sub-scores) feature subsets.

The full model yielded modest classification performance (AUC = 0.515, balanced accuracy = 0.508, Cohen's kappa = 0.016). DTI-only and Vascular Burden Score-only feature subsets yielded AUCs of 0.571 and 0.282 respectively. Permutation variable importance was dominated by the three nigral diffusion metrics (RD = 0.213, FA = 0.144, MD = 0.119), with smaller but non-zero contributions from the Vascular Burden Score microvascular (0.041) and macrovascular (0.020) sub-scores. The pattern is consistent with Vascular Burden Score capturing variance that is largely orthogonal to nigral diffusion microstructure rather than redundant with it. Given the small imaging subset and modest overall classification performance, this exploratory analysis is reported as hypothesis-generating; replication in larger imaging-phenotyped cohorts is required. Detailed results are in Supplementary Table S6, with the permutation importance ranking shown in Supplementary Figure S2.

### Supplementary Figure S1

**Vascular factor prevalence by diabetes mellitus/prediabetes status across the PPMI diagnostic cohorts (N = 413).**

Prevalence of each of the seven vascular risk factors (hypertension, hyperlipidemia, obstructive sleep apnea, obesity, stroke/TIA, coronary artery disease, atrial fibrillation) shown by diabetes mellitus/prediabetes group across the full sample. Microvascular factors (hypertension, hyperlipidemia, sleep apnea, obesity) show consistent elevation in the diabetes mellitus/prediabetes group across cohorts, while macrovascular factors (stroke/TIA, coronary artery disease, atrial fibrillation) show no consistent group difference. This supplementary figure visualizes the data underlying the per-factor adjusted prevalence ratios reported in Table 2 of the main manuscript.

**Supplementary Figure S2**

**Random Forest Permutation Variable Importance (n = 50).**

Permutation variable importance (mean across 200 iterations) for the five features entered into the random forest classifier distinguishing healthy controls (n = 22) from clinically diagnosed Parkinson's disease participants (n = 28). Features are ranked by importance. The three nigral diffusion tensor imaging metrics (radial diffusivity, fractional anisotropy, mean diffusivity) dominate the importance ranking, with smaller but non-zero contributions from the Vascular Burden Score microvascular and macrovascular sub-scores. Full model AUC = 0.515; DTI-only feature subset (FA, MD, RD) AUC = 0.571; Vascular Burden Score-only feature subset AUC = 0.282. The pattern is consistent with Vascular Burden Score components capturing variance that is largely orthogonal to nigral diffusion microstructure rather than redundant with it. See Supplementary Analysis S2 for full methodology and Supplementary Table S6 for detailed results.
