## Supplementary Figures for "Vascular Phenotyping in Parkinson’s Disease: Diabetes Mellitus Operationalizes a Microvascular Metabolic Syndrome Cluster Across PPMI Diagnostic Cohorts"

### Slide 1
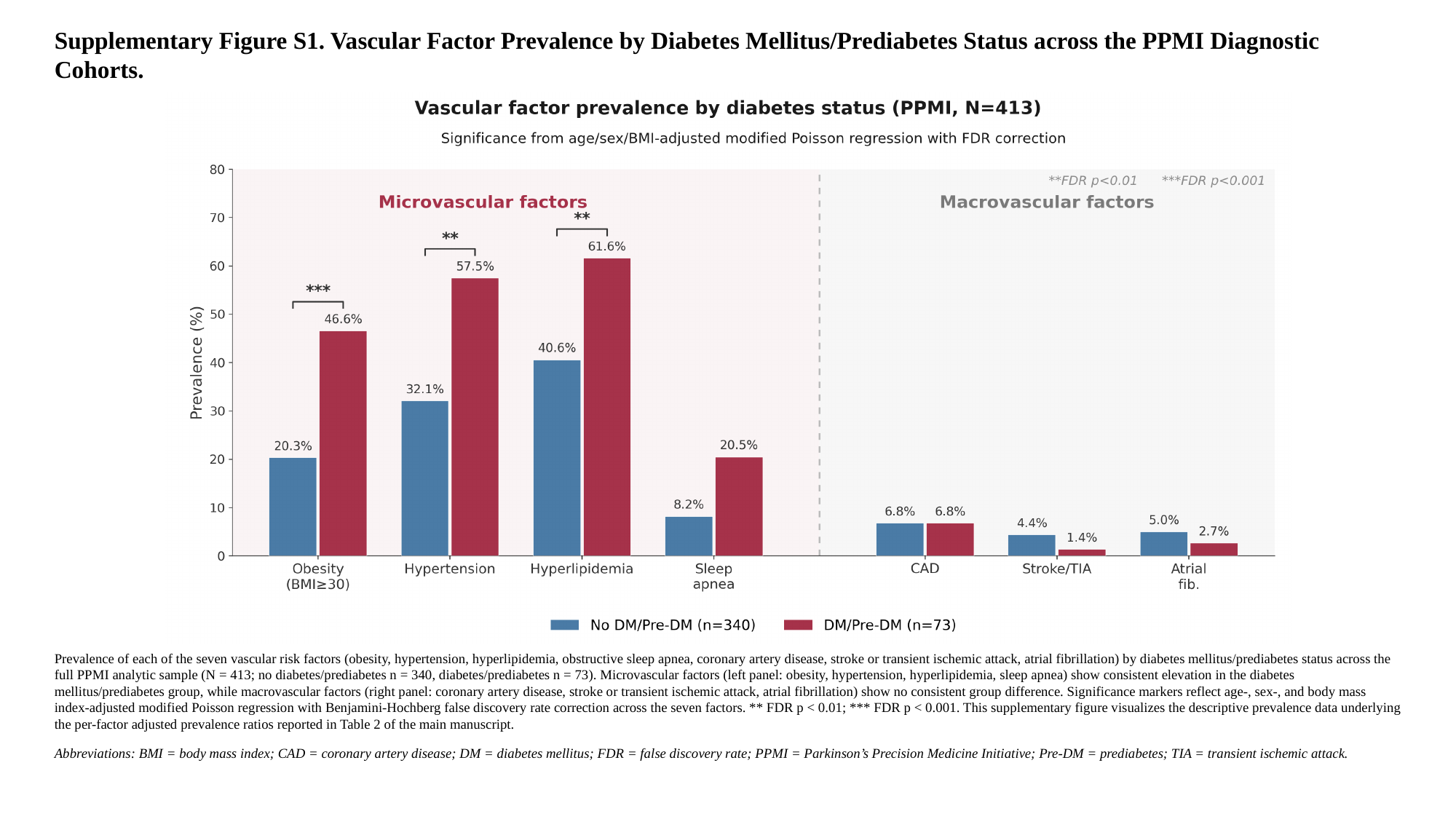

Supplementary Figure S1. Vascular Factor Prevalence by Diabetes Mellitus/Prediabetes Status across the PPMI Diagnostic Cohorts.
Prevalence of each of the seven vascular risk factors (obesity, hypertension, hyperlipidemia, obstructive sleep apnea, coronary artery disease, stroke or transient ischemic attack, atrial fibrillation) by diabetes mellitus/prediabetes status across the full PPMI analytic sample (N = 413; no diabetes/prediabetes n = 340, diabetes/prediabetes n = 73). Microvascular factors (left panel: obesity, hypertension, hyperlipidemia, sleep apnea) show consistent elevation in the diabetes mellitus/prediabetes group, while macrovascular factors (right panel: coronary artery disease, stroke or transient ischemic attack, atrial fibrillation) show no consistent group difference. Significance markers reflect age-, sex-, and body mass index-adjusted modified Poisson regression with Benjamini-Hochberg false discovery rate correction across the seven factors. ** FDR p < 0.01; *** FDR p < 0.001. This supplementary figure visualizes the descriptive prevalence data underlying the per-factor adjusted prevalence ratios reported in Table 2 of the main manuscript.
Abbreviations: BMI = body mass index; CAD = coronary artery disease; DM = diabetes mellitus; FDR = false discovery rate; PPMI = Parkinson’s Precision Medicine Initiative; Pre-DM = prediabetes; TIA = transient ischemic attack.

### Slide 2
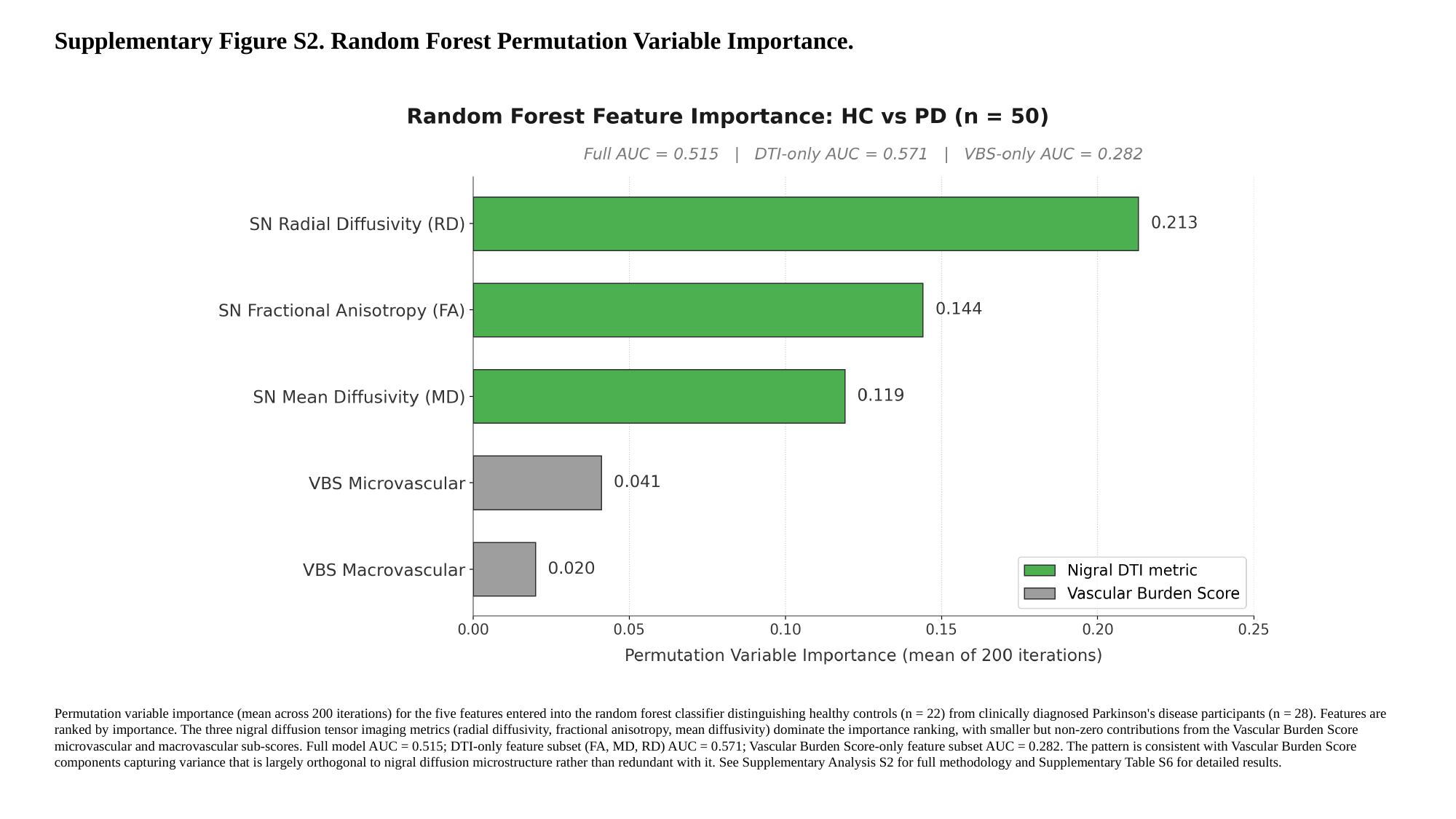

Supplementary Figure S2. Random Forest Permutation Variable Importance.
Permutation variable importance (mean across 200 iterations) for the five features entered into the random forest classifier distinguishing healthy controls (n = 22) from clinically diagnosed Parkinson's disease participants (n = 28). Features are ranked by importance. The three nigral diffusion tensor imaging metrics (radial diffusivity, fractional anisotropy, mean diffusivity) dominate the importance ranking, with smaller but non-zero contributions from the Vascular Burden Score microvascular and macrovascular sub-scores. Full model AUC = 0.515; DTI-only feature subset (FA, MD, RD) AUC = 0.571; Vascular Burden Score-only feature subset AUC = 0.282. The pattern is consistent with Vascular Burden Score components capturing variance that is largely orthogonal to nigral diffusion microstructure rather than redundant with it. See Supplementary Analysis S2 for full methodology and Supplementary Table S6 for detailed results.
Abbreviations: AUC = area under the receiver operating characteristic curve; DTI = diffusion tensor imaging; FA = fractional anisotropy; HC = healthy control; MD = mean diffusivity; PD = Parkinson's disease; PPMI = Parkinson's Progression Markers Initiative; RD = radial diffusivity; SN = substantia nigra; VBS = Vascular Burden Score.
